# Improving Care for Amyotrophic Lateral Sclerosis with Artificial Intelligence and Affective Computing

**DOI:** 10.1101/2024.10.07.24314892

**Authors:** Marc Garbey, Quentin Lesport, Gülşen Öztosun, Veda Ghodasara, Henry J. Kaminski, Elham Bayat

## Abstract

**Background:** Patients with ALS often face difficulties expressing emotions due to impairments in facial expression, speech, body language, and cognitive function. This study aimed to develop non-invasive AI tools to detect and quantify emotional responsiveness in ALS patients, providing objective insights. Improved understanding of emotional responses could enhance patient-provider communication, telemedicine effectiveness, and clinical trial outcome measures.

**Methods:** In this pilot study, fourteen patients with ALS had audio recordings performed during routine clinic visits while wearing a wireless pulse oximeter. Emotion-triggering questions related to symptom progression, breathing, mobility, feeding tube, and financial burden were randomly asked. The same questions were posed in separate psychiatric evaluations. Natural language processing (NLP) was used to analyze transcriptions, topic classifications, sentiment, and emotional states, combining pulse and speech data. AI-generated reports summarized the findings.

**Results:** Pulse alterations consistent with emotional arousal were identified, with longer consultations and positive communication reducing pulse fluctuations. Financial concerns triggered the strongest emotional response, while discussions about breathing, mobility, and feeding tube increased anxiety. AI-generated reports prioritized patient concerns and streamlined documentation for providers.

**Conclusions:** This study introduces a novel approach to linking pulse and speech analysis to evaluate emotional responses in ALS patients. AI and affective computing provide valuable insights into emotional states and disease progression, with potential applications for other neurological disorders. This approach could augment clinical trial outcomes by offering a more comprehensive view of patient well-being.

## 1. Introduction

Amyotrophic lateral sclerosis (ALS) is a progressive neurodegenerative disease that significantly impacts a patient’s ability to communicate verbally, through facial reactions, and general body language. These make patient care [1] particularly challenging for all healthcare professionals. Maximizing the information gained during the brief, episodic clinic visits and supporting positive adaptive behaviors as the disease progresses requires appreciation of the patient’s emotional responses from the multi-disciplinary care team [2] [3]. The psychological state of ALS patients when formally assessed shows great variability and can be better than expected by the healthcare practitioner; [4] however, detailed psychological assessments are often not part of routine visits, even in comprehensive care clinics, and even less so for patients in resource-poor environments. In addition, even for the most experienced physician, non-verbal communication of emotions may be difficult to interpret.

Typically, patients will have periodic clinic visits with interval progression of weakness, and the clinician needs to anticipate the need to discuss key treatment choices, keeping in mind that each patient is unique in their coping skills and particular priorities for their lives. For instance, difficult topics such as tube feeding, and respiratory support are all necessary. Similarly, use of wheelchairs, bed device assistance, communication devices, and respiratory support designed to preserve the patient’s autonomy, communication capabilities, and metabolic functions need to be explained. Consideration of financial and insurance coverage are critical, adding to the huge burden on patients and care-givers.

Often, the patient is unprepared to have emotionally charged discussions, and yet keeping their morale up is an essential component to enhance quality of life and assure effective collaboration with care team. Providing empathy during a clinical consultation requires that the physician reads the patient and evaluates their emotional state through verbal and non-verbal communication [3]. A common challenge is balancing a patient’s subjective experiences with their objective medical conditions to provide personalized care. When patients either deny their symptoms or overreact to them, a lack of understanding and anticipation can exacerbate the potential for a poor therapeutic relationship with great frustration for the physician, care team, care-givers, and the patient.

Nonverbal communication of emotions in patients can be assessed through computer vision and artificial intelligence (AI) approaches [4,5,6,7]. Rosalind Picard recognized the potential of affective computing [8] with analysis of emotion in speech and written text making dramatic progress recently [9,10,11]. While many of these techniques to assess a patient’s emotional state from video, speech, or text are presented as versatile and scalable by computer scientists, they underestimate the complexity of patient care and specific needs for care of patients with motor impairment [12,13]. This is particularly true in ALS [14] due to muscle weakness with impairment of facial expression, speech [15,16], and body language, as well as cognitive and behavioral changes. ALS patients may have emotional perception deficits and react poorly to nonverbal communication of empathy by others, including their physicians [15,17].

We propose an AI methodology that includes emotional computing to enhance the quality of listening to the patient during a standard clinical encounter and integrate with objective physical examination and supplement information from the patient. To make those results implementable, the assessment can provide automatic and systematic report documentation intended to improve understanding of patient needs across multi-disciplinary team. The psychiatrist, speech therapist, social worker, nutritionist and physical therapist, for example, can benefit from a real time objective report that quantitatively assesses the patient’s priorities and concerns that may not be typically appreciated.

## 2. Methods

### 2.1 Human Subjects

Fourteen subjects were recruited from the multidisciplinary ALS clinic at George Washington University. Subjects had clinical and electrophysiologic confirmation of ALS. Subjects were older than 18 years, with no other unstable medical condition that would pose a risk to participation according to the examining physician’s judgment or had cognitive impairment, dementia, or a psychiatric illness prior to the diagnosis of ALS. Patients at a terminal stage of the disease that necessitated tube feeding, were unable walk, or had significant dysarthria were excluded from the study. All patients provided written consent, and the study was approved by the institutional review board of George Washington University.

### 2.2 Study Protocol

Patients completed the ALS Functional Rating Scale (ALS-FRS) prior to the clinical examination [18]. Subjects participated in the study during their routine clinic visit with no prolongation of the typical appointment time. A wireless pulse oximeter (OxyU Vibeat, Shenzhen China) was worn by the patients throughout the visit to record their heart rate and oxygen saturation. The patients’ and the neurologist’s voices were recorded using a Zoom-H1n audio recorder during the entire visit except for the psychiatry session. After the psychiatry session, the psychiatrist wrote a report describing the patients’ mood and emotional state.

Subjects were asked to respond to three questions asked by the neurologist and psychiatrist to assess emotional response (Table 1). The psychiatrist recorded the timepoints when these questions were asked as the patients’ voices were not recorded during this session.

**Table 1.**
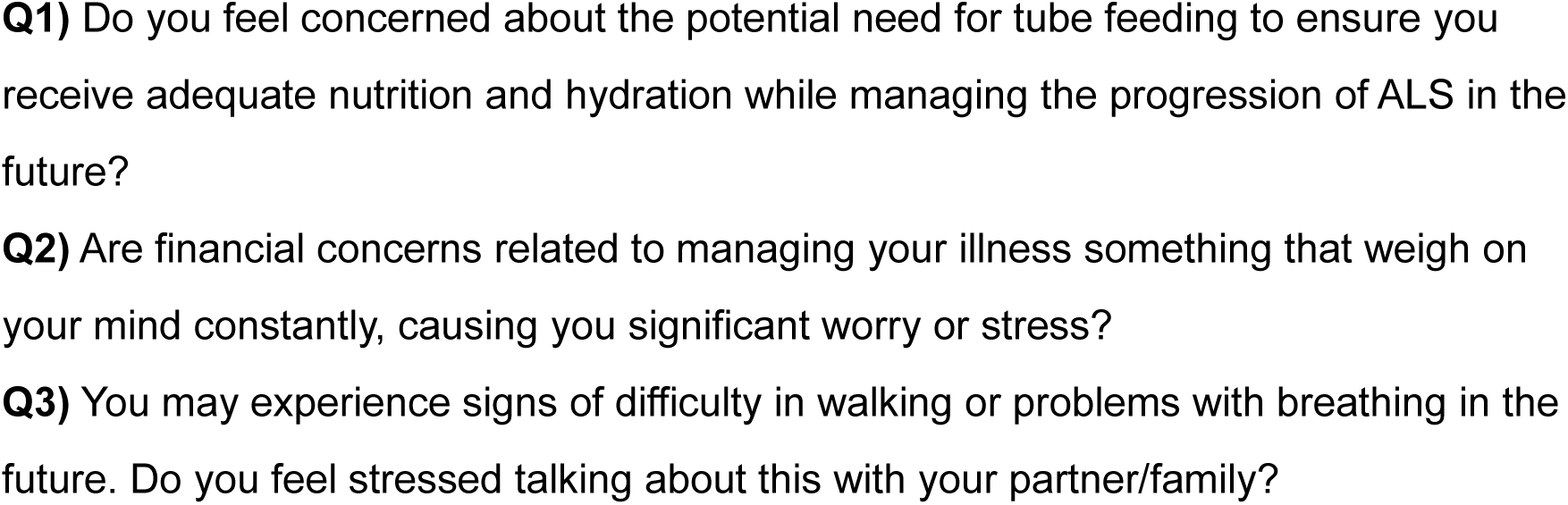
Emotional Response Questions.

The pulse oximeter continuously monitored oxygen levels, pulse rate, and body movement during the session with the neurologist and subsequent psychiatry session (typically consecutive, lasting 20-30 minutes each). Pulse data was collected every 3 seconds. The manufacturer reports a pulse oximeter range of 30-250 bpm with ±2 bpm or ±2% precision (whichever is greater). The OxyU Vibeat is not FDA-approved but is comfortable and unobtrusive for patients during examinations. We compared pulse readings from the OxyU Vibeat to an FDA-cleared CMI (PC-66L) oximeter, demonstrating consistency (data not shown). A high-fidelity Zoom-H1n audio recorder (Zoom Corp, NY) captured audio during the clinical session only. This recorder features stereo X/Y 90° microphones, handles up to 120 dB SPL and supports various audio formats.

### 2.3 Natural Language Processing

We leveraged an AI transcription tool, AssemblyAI [19], to generate transcripts of the recorded clinical sessions. These transcripts included timestamps for each word, confidence scores, and speaker identification. The transcripts were segmented into sentences categorized by speaker (doctor, patient, or companion) and separated questions from statements. To facilitate topic classification, all transcripts were combined into a single file. We then built a dictionary of key terms for classifying sentences into six categories related to nutrition, breathing, walking, speech, medication, finance, and miscellaneous. This dictionary development employed a combination of manual extraction and generative language processing using ChatGPT. The dictionary has the capability to be refined and enhanced to minimize Natural Language Processing (NLP) misclassifications. Our analysis yielded several key metrics, including the relative weight of conversation topics and subject participation levels.

### 2.4 Affective Computing

To assess patient emotional arousal, we employed the following techniques:

Physiological Data: We analyzed pulse rate focusing on rapid changes that might indicate anxiety [20,21]. We filtered out noise and assessed localized variations in the time interval on the order of 10 seconds exceeding 4 pulses per minute to ensure reliable detection and referred to these localized changes as a “pulse event”. Additionally, we ensured data validity by excluding measurements potentially affected by patient movement. We also computed heartbeat variation trend over the period of the examination using a least square linear fit of the signal [22]. Increasing trends could suggest that the patient is becoming more nervous. Alternatively, a decreasing trend may point out to the patient becoming more relaxed. We could use this indicator, only if the least square fit is accurate.

Sentiment Analysis: We used the Valence Aware Dictionary and Sentiment Reasoner (VADER) sentiment lexicon [23] to analyze the transcript text and identify sections with the most negative sentiment, potentially reflecting emotional distress. This sentiment analysis approach can be further refined and specialized to ALS patients as a larger data set is analyzed.

Voice Analysis: We extracted features from the patient’s speech, including pitch and loudness. We compensated for potential fatigue by removing long-term trends in decrease of the loudness signal. We specifically identified sudden increases in loudness, which could indicate frustration or emphasis. While pitch was included in our analysis, it may prove to be unreliable due to potential ALS-related speech impairments.

Speech Rate: We calculated the time interval between words, as speaking speed may be influenced by emotion. The analysis generated a dozen features reflecting patient and doctor communication patterns. We used these features to analyze the entire clinical session or focus on specific triggers, such as the pre-defined questions in the protocol.

We extracted sentences corresponding to significant events identified through the above methods. The sentences (Table 1) were combined into a “condensed transcript” annotated with timestamps for each event. This condensed transcript was typically about 20% of the original transcript length and is well-suited for further analysis.

### 2.5 Metrics

We conducted a correlation analysis to investigate the relationship between various human factors and results of AI analysis. We assessed correlation between sentiment analysis metrics, duration of the clinical examination, percentage of the duration physician talking, patient self-assessment feedback, and other relevant variables. For each analysis we determined a triplet of statistical results: 1) Pearson correlation coefficient, indicating the strength and direction of the linear relationship between the variables. 2) P-value, representing the probability of observing a correlation as extreme as the calculated one, assuming there is no true correlation. 3) Sample size, indicating the number of observations used in the analysis. Additionally, we included a graphical visualization to aid in understanding the correlation. The visualizations were scatter plots, showing the relationship between the two variables and the overall trend. While these metrics can provide valuable insights, they do not establish causality.

### 2.6 Generative Language Processing Integration

We employed Generative Language Processing (GLP) techniques in conjunction with our AI methods for speech analysis to produce verifiable results and guide the language generation process towards specific questions or inputs, enhancing the robustness of the observations. For example, we utilized ChatGPT to generate a dictionary of keywords for each topic using all transcripts from subject assessments. We then manually reviewed and validated this dictionary. Our approach focused on the condensed transcript which is a subset of the full transcript containing key moments identified through affective computing methods described above. We hypothesized that this condensed transcript captures the most relevant aspects of the therapeutic conversation. Utilizing ChatGPT, we requested the following analyses:

2.6.1 Ranking Patient Concerns: Based on the condensed transcript, ChatGPT ranked the patient’s level of concern for each topic. This ranking could subsequently be compared with the psychiatrist’s report for validation.
2.6.2 Graphical Representation: A user-friendly pie chart was generated to visually represent the ranked concerns.
2.6.3 Medical Report Generation: ChatGPT was asked to draft a concise report for each topic and a summary of the entire conversation based on the condensed report and the ranking.

### 2.7 Software Architecture

Our software utilizes a modular architecture, combining MATLAB, Python, and the ChatGPT API. MATLAB serves as the core platform, handling most signal and NLP analysis tasks. Python libraries or the ChatGPT API can be called directly from the MATLAB code. This modular approach allows for flexibility and potential future integration of additional tools. The software runs in approximately 5 minutes per patient, enabling quick analysis to support real-time collaboration within the multidisciplinary care team. This approach offers the potential to automatically generate reports using LaTeX [https://www.latex-project.org/] or similar tools.

While currently not implemented, integration of a report with an electronic medical record for report and data upload is feasible. This functionality would facilitate longitudinal monitoring of patients.

## 3. Results

### 3.1 Patient Characteristics

Table 2 shows study population characteristics. All subjects except 4 had spinal onset ALS and were early in the disease course (Figure 1).

**Figure 1.**
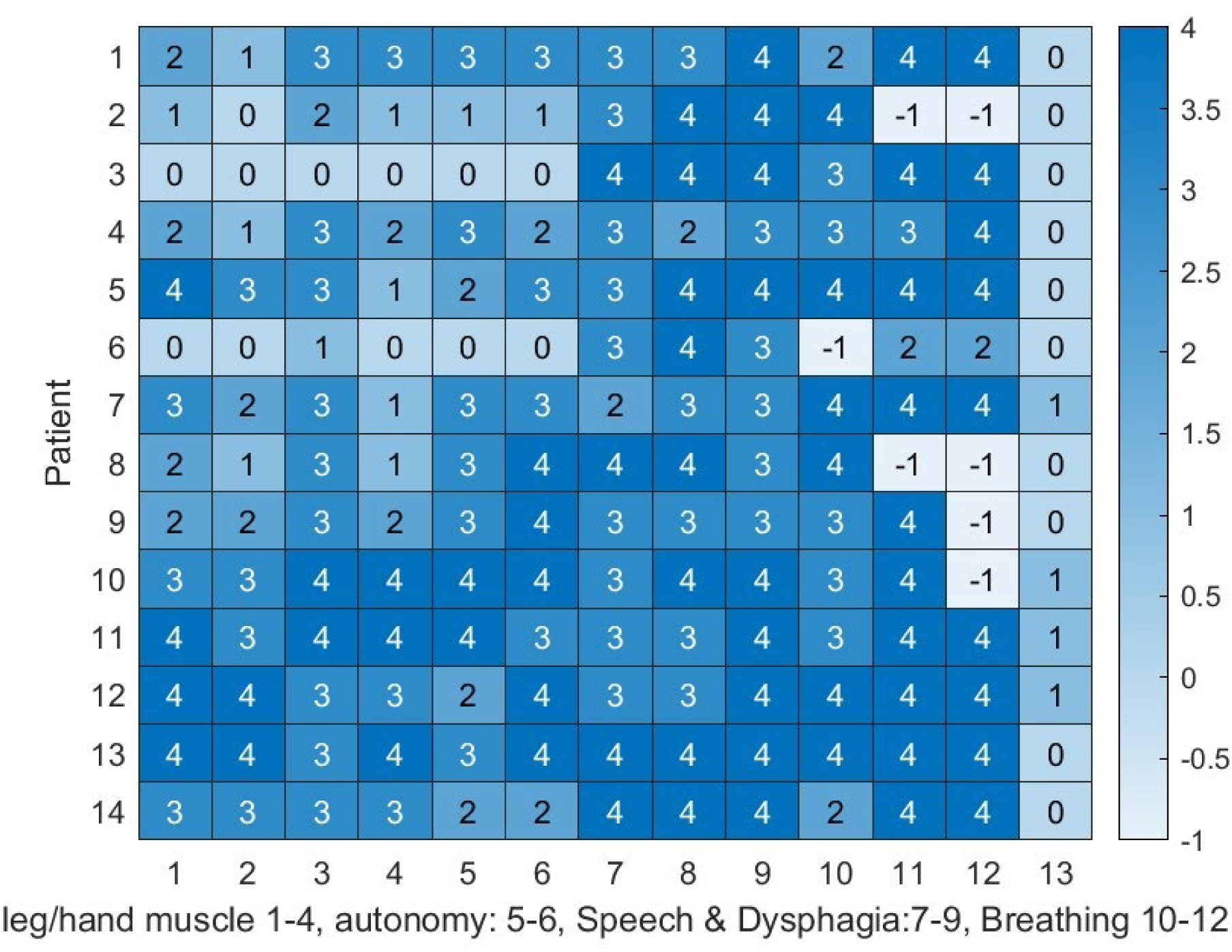
Heatmap summarizing the ALS-FRS scores for all subjects, visually representing the disease severity and progression, with scores (4 = normal, 0 = the worst, -1=N/A). The order of entries corresponds to the ALSFRS categories: leg muscle weakness minutes, hand muscle weakness, autonomy (dressing, hygiene, turning in bed), speech/salivation/dysphagia, and breathing (dyspnea, orthopnea, difficulty breathing). The last column is 0 for spinal ALS onset and 1 for bulbar onset.

**Table 2:** Patient Demographics and Clinical Characteristics.

| Patient | Age Range | Race | Gender | Clinical Presentation at Onset | Months Since Onset |
| --- | --- | --- | --- | --- | --- |
| 1 | 65-74 | White | Male | Spinal | 10-20 |
| 2 | 75+ | White | Male | Spinal | 21-30 |
| 3 | 75+ | Non-White | Male | Spinal | 50+ |
| 4 | 75+ | White | Female | Spinal | 10-20 |
| 5 | 75+ | Non-White | Male | Spinal | 10-20 |
| 6 | 65-74 | White | Male | Spinal | 10-20 |
| 7 | 36-49 | White | Female | Bulbar | 21-30 |
| 8 | 50-64 | White | Female | Spinal | 10-20 |
| 9 | 65-74 | White | Female | Spinal | 10-20 |
| 10 | 65-74 | White | Female | Bulbar | 10-20 |
| 11 | 75+ | Non-White | Female | Bulbar | 10-20 |
| 12 | 50-64 | White | Female | Bulbar | 10-20 |
| 13 | 65-74 | White | Male | Spinal | 10-20 |

### 3.2 Oximeter Use

At the start of the session, each patient indicated the oximeter was comfortable after placement. However, the sensor did not provide a pulse signal for an extended period during the examination for subjects 4, 7, and 8. This oximeter appeared to be too sensitive to finger size, skin condition and sensor placement for these individuals and one inadvertently removed the probe.

### 3.3 Response to Questions

Figure 2 explores patient responses to questions about nutrition and breathing during both the neurology and psychiatry sessions. There was good correlation between the assessments. Questions 1 and 2 were correlated between the two sessions (Table 3) but patient responses would vary between the two sessions, which were typically 30 minutes apart suggesting that clinicians may need to address these topics repeatedly to ensure patient understanding. One potential explanation for some of the variation is early cognitive impairment. We found during the examination of Patient 11 that they were developing symptoms of frontotemporal dementia, and therefore could have had difficulty processing the questions.

**Figure 2.**
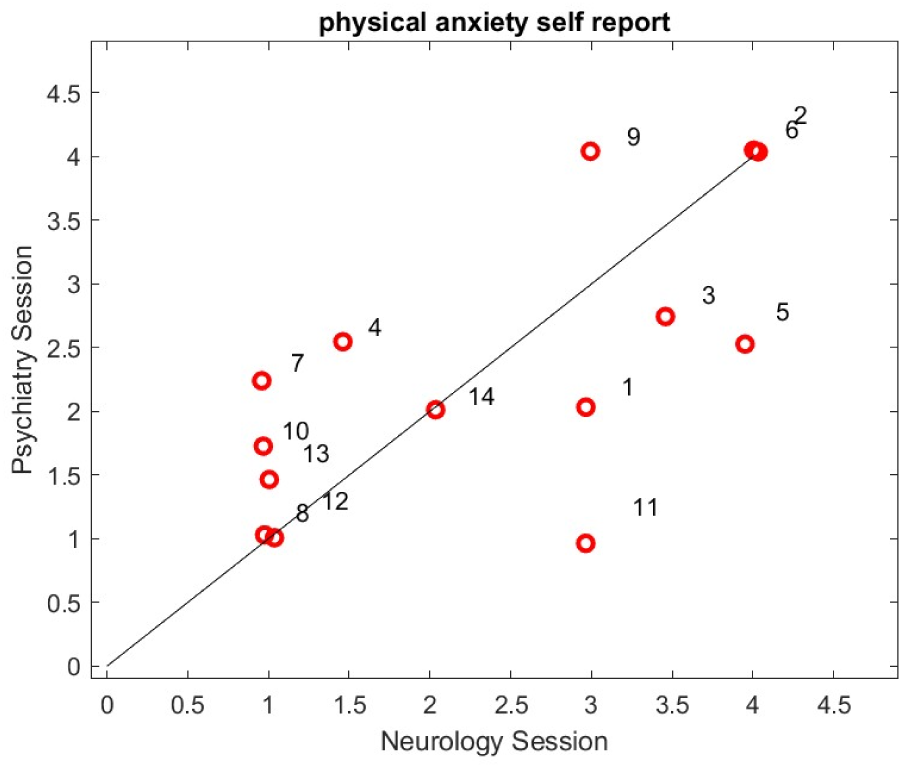
Patient responses to questions about nutrition and breathing during both the neurology and psychiatry sessions. The horizontal axis represents the average score from the neurology session, and the vertical axis represents the average score from the psychiatry session. Each red circle corresponds to a patient. Score values: 4: serious concern, 3: moderate concern, 2: mild concern, 1: no concern

**Table 3.** Correlation assessment of questions to patient responses.

| Question: | r | p | S | Visualization |
| --- | --- | --- | --- | --- |
| 1. Is physical anxiety self-report during clinic consistent? | 0.66 | 0.01 | 14 | Figure 2 |
| 2. Is mean sentiment score communicative? | 0.69 | 0.0061 | 14 | Figure 3 |
| 3. Is pulse trend related to duration of clinical examination? | 0.02 | 0.94 | 11 | Figure 5A |
| 4. Is patient stress related to the time the doctor speaks? | 0.65 | 0.04 | 10 | Figure 4 |

Regarding the financial question, patients clearly understood the question, which was typically only asked once during the clinical session. Patient responses showed less variability between sessions, suggesting a consistent level of concern. Financial difficulties can be a significant stressor, and some patients may be hesitant to answer due to perceived unfairness from the healthcare system. Regardless, the medical team has a responsibility to assess the financial impact of the disease and offer guidance on available resources.

### 3.4 Communication and Patient Anxiety

We investigated separately the potential link between longer consultations or positive communication and patient anxiety. Figure 3 depicts the correlation between the sentiment analysis of patient and neurologist speech during the examination. The results suggest that positive/negative sentiments can be contagious during communication, although the direction of influence is unclear.

**Figure 3.**
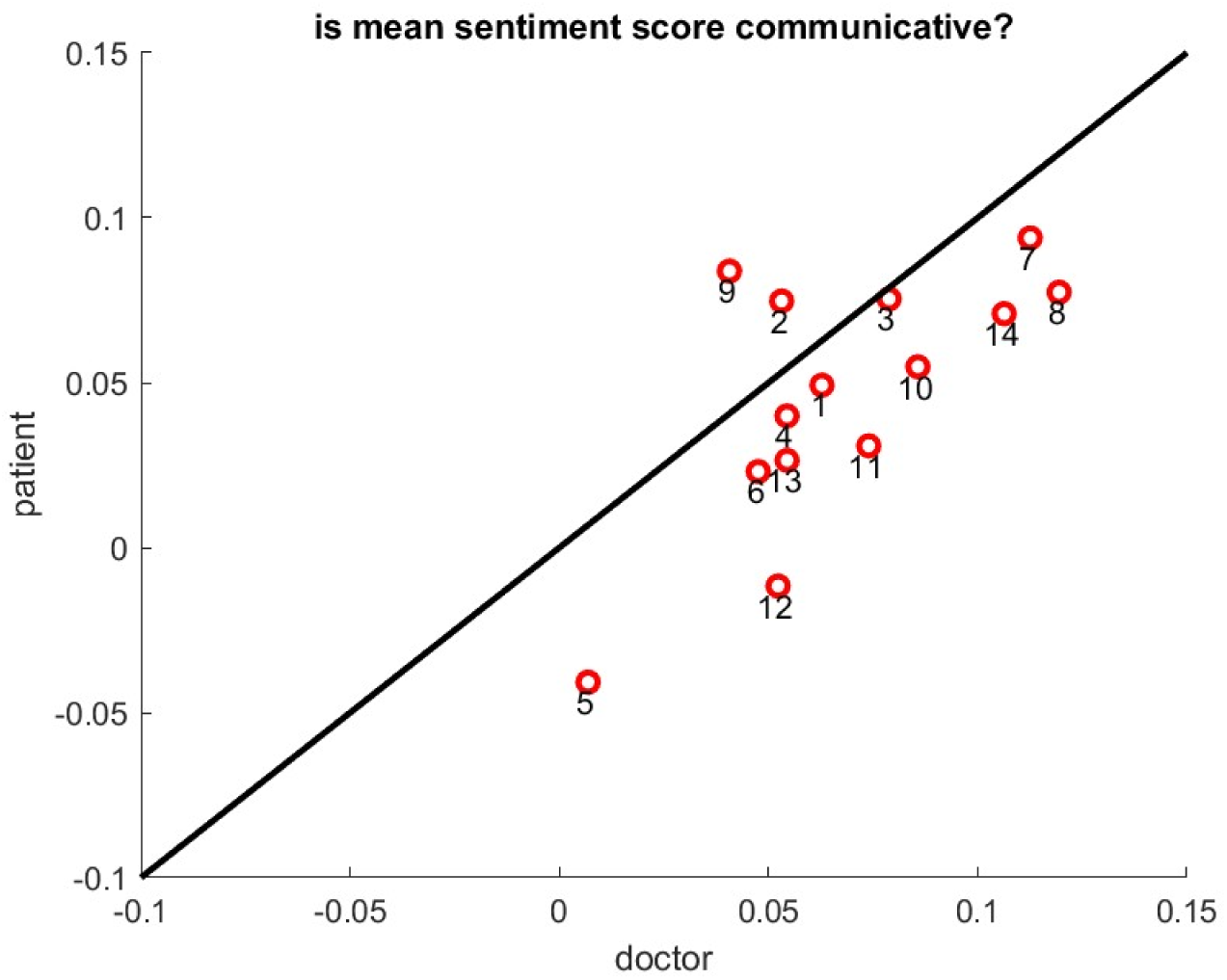
Scatter plot represents the average sentiment analysis score provided by the VADER algorithm of the patient’s speech versus the neurologist’s speech during the clinical consultation. Each red circle corresponds to a subject.

The second row of Table 3 indicates a quantitative assessment of the correlation. The neurologist, who has extensive experience in care of patients with ALS, typically speaks for about 70% of the consultation which may indicate that she can positively influence patient mood.

Figure 4 complements this finding by showing a connection between the time the neurologist speaks and a decrease or increase in patient heart rate variability, indicating a calmer state or more anxious state. The correlation (Table 3 second row) suggests that positive discussions with the neurologist may help patients relax during the consultation. The duration of the session did not correlate with heart rate trends, indicating that fatigue of the patient was not a factor (Table 3 third row). Again, patient 11 appears as an outlier, potentially due to an undiagnosed early-stage dementia limiting her ability to react to the doctor’s explanations. This single patient suggests the potential utility of using heart rate variability to contribute to the detection of early cognitive impairment during the course of ALS.

**Figure 4.**
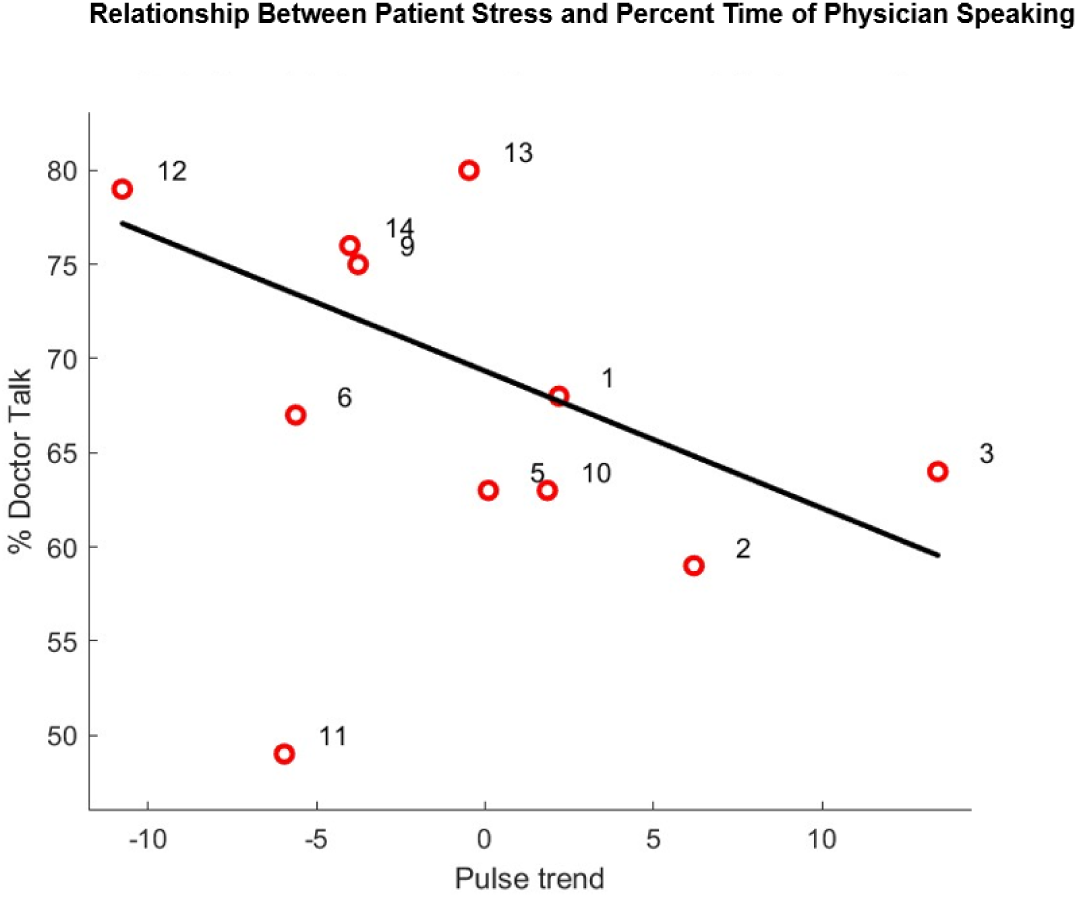
Scatter plot of the horizontal coordinate is the slope of the least square linear of the pulse during the neurology session. The vertical coordinate is the relative percentage of time the neurologist is speaking.

### 3.5 Digital Markers of Anxiety

We examined the relationship between heart rate variability and patient anxiety during the clinical encounter. Figure 5 presents various anxiety measures: the accumulated number of Delta Pulse Events (DPE) normalized by the total time elapsed during the session, denoted DPE, as well as physician assessments on stress and mood. We could only compute all three metrics for 8 subjects. The number is too small for the assessment of stress and mood to support a correlation analysis as in Table 2; however, qualitative observations can be made. 1) Patients 1 and 6 expressed high stress levels with large DPE indexes. 2) Patient 2 declares that he was overwhelmed and has the lowest DPE index. 3) Patient 11 has early FTD based on clinical evaluation and has a moderate DPE index. A larger population and detailed psychiatric evaluation to obtain quantitative analysis will be required.

**Figure 5:**
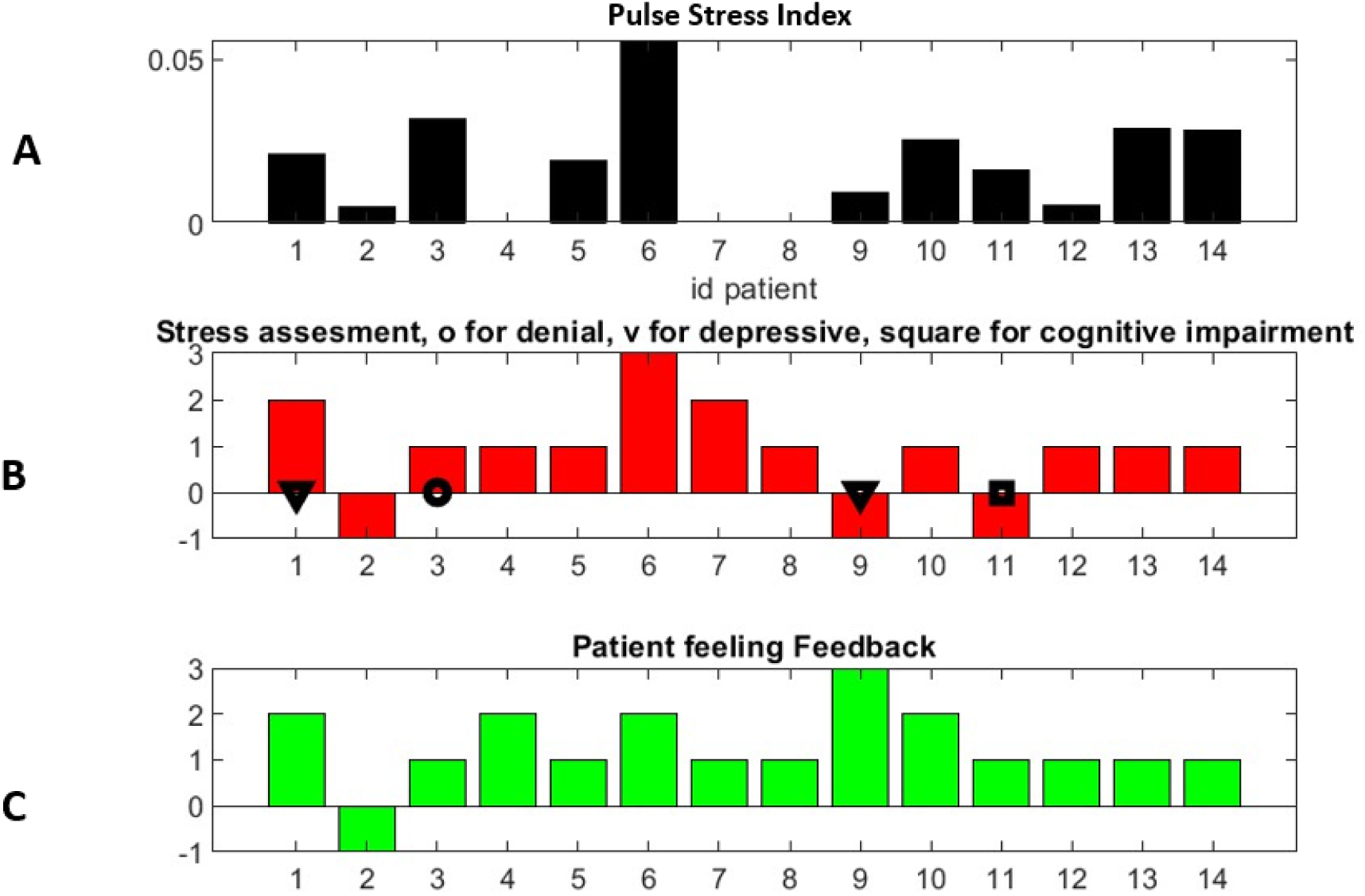
Figure 5A shows the accumulated number of delta pulse events normalized by the total time elapsed during the session.

Figures 5B, 5C attempt to quantify the psychiatry report. Patient was asked to report whether they were very good (1), average (2) or not good (3).

Stress level when explicitly reported goes from 1 (low) to 3 (very high). -1 indicates information is missing. Circle and Triangle are annotations for state of denial or depression. Square suggests cognitive impairment on figure 5B based on neurologist’s determination

Pulse is not available for patients 4-7-8. Stress evaluation is not available for patients 2, 9, 11

Figure 6 demonstrates a clear pattern for the question related to financial impact. In principle, this question was asked only once and did not lead to extended discussions, unlike the other two questions, that aimed to prompt conversation about specific symptoms.

**Figure 6:**
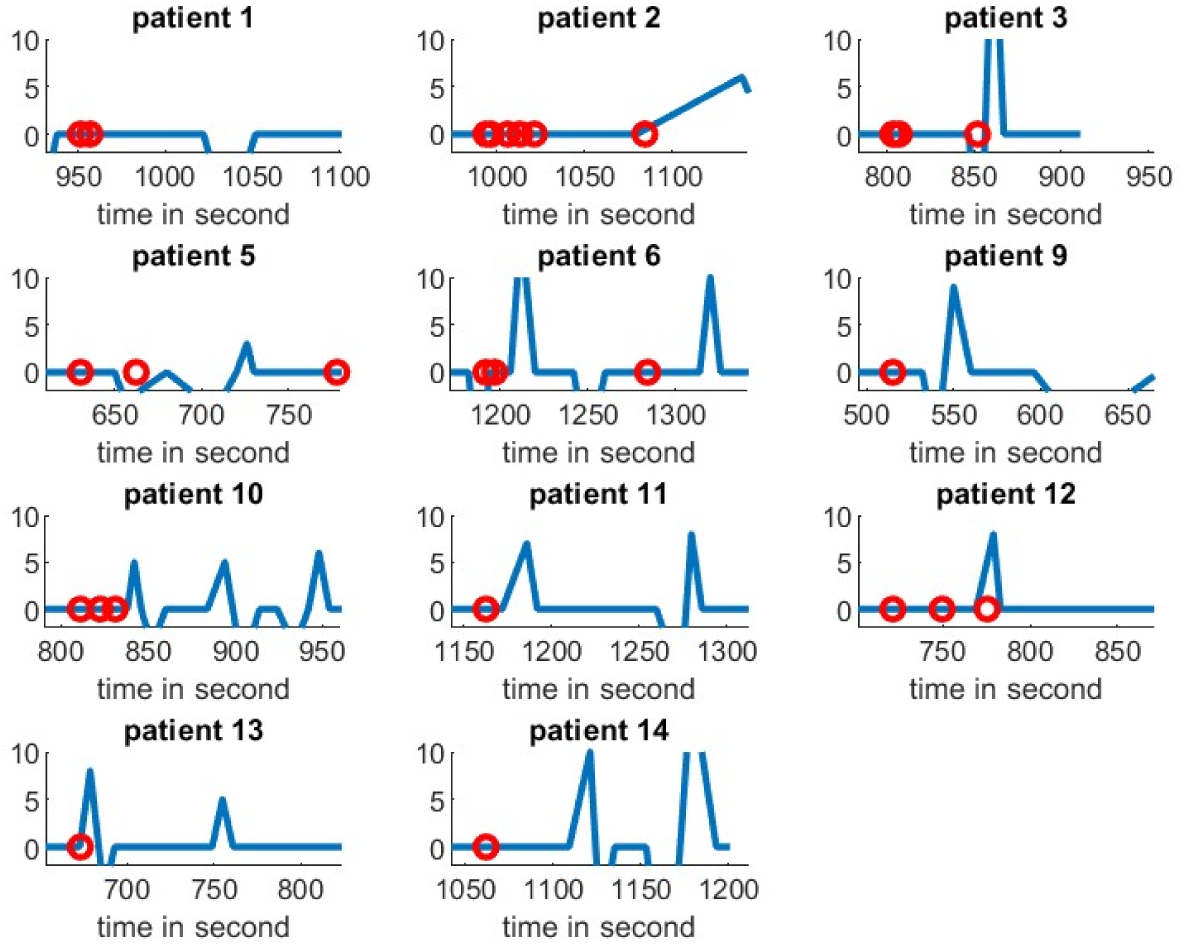
Pulse variation that is occurring during response to financial question. Red circle indicates only when the finance questions are discussed during the full session. Patients 1, 5, 10, 12 and 14 consistently reported no financial concerns. Our digital markers corroborated this for patients 1,5 and 14, but not for patients 10 and 12. Patient concerns on finance aligns with the findings for patients 2, 3, 6, 9 and 13. Interestingly, patient 9 changed his mind, reporting no concern during the clinical session but high concern during the psychiatry session. Patient 11 refused to answer the financial question but exhibited a high response. Our digital marker appeared to capture his true anxiety level already during the neurology session. The sample size is insufficient for statistical analysis.

The annotations (red circle) indicate when the questions related to finance were asked. If a pulse peak in Figure 6 follows closely the red circle time stamp, one considers that the patient reacts with a pulse increase to the question. The higher peak indicates a greater the reaction.

### 3.6 Clinical Actionable Outputs

To facilitate clinical decision-making, the AI algorithm automatically generated a report summarizing the patient’s key concerns, along with a corresponding visual representation (Figure 7).

**Figure 7:**
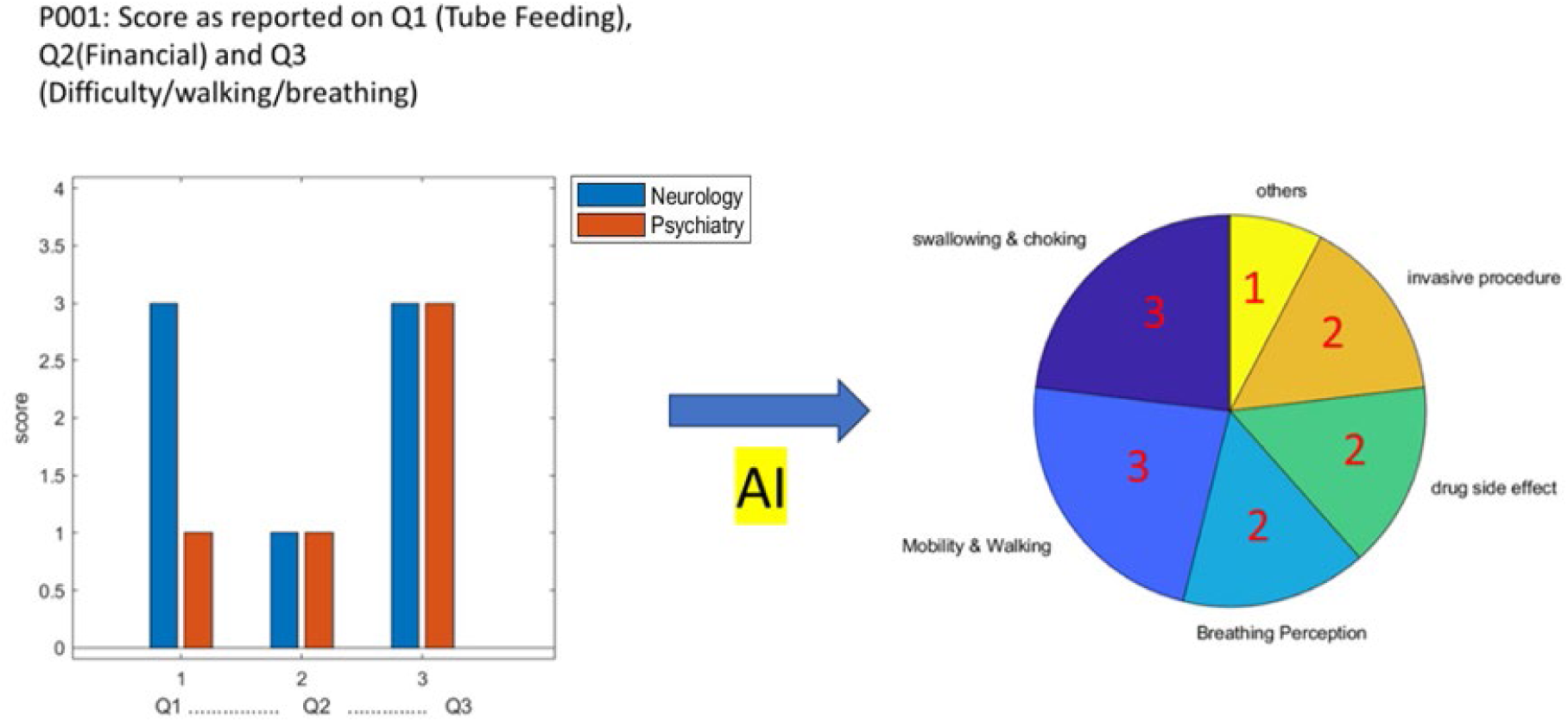
Example of quick graphic overview of patient concerned ranked by categories generated automatically by the AI algorithm.

Appendix A provides an example for patient 1. This output can be customized to meet the clinician’s specific needs. Annotations can be added through a user-friendly interface like a word processing environment. Due to this process being entirely automated and rapid, these reports can be readily available for the multidisciplinary team caring for the patient on the same day, enhancing overall patient care efficiency.

## 4. Discussion

Assessment of emotional responses during clinic visits of patients with ALS are challenging. For example, complex topics are often broached, such as the decision to initiate a feeding tube, with patients perceiving the benefits and drawbacks quite differently [24]. While professional experience can guide physicians, it does not guarantee comprehension of a patient’s unspoken concerns. Further, assessment of a patient’s emotional needs is challenging for even experienced clinicians. Non-invasive assessment of a patient’s emotional response could be a valuable adjunct to care. [25, 26] A rational assessment of emotional state during these discussions could be a vital guide to optimal care management, including the best combination of invasive respiratory devices, comfort medications, and psychological support. Our primary target in this clinical study was to evaluate patients’ emotional feedback on specific topics. Our secondary target was to identify emotional issues from the patient perspective that may have not been anticipated by the physician.

We expanded upon the typical assessments by a physician in asking, “Are financial concerns related to managing your illness a constant source of worry or stress?” This question is crucial for understanding emotional arousal in ALS patients. Given the increasing complexity of healthcare costs and insurance reimbursement, the multidisciplinary clinical team should not overlook the significant impact of financial factors on patient well-being. While this question raises ethical considerations beyond the scope of this paper, it highlights the importance of addressing financial concerns in clinical care.

Our computational method to identify emotional surges and label them was based on the following basic principles. 1) Heart rate escalation is a well-established measure of emotional arousal [20,27] and is nearly universally found across species [21]. Jauniaux and colleagues [20] showed that rapid variation of heart rate is correlated to emotional regulation, in contrast to other studies [28,29] that examine the long-term effect of mood on heart rate measures. The heart rate appears not associated with depression scales in long-term studies [28] and does not differentiate anger and fear from neutral emotional state [29].

Further one expects that pulse varies with body motion, especially for patients who have muscular weakness affecting their respiratory functions. One can use the metric of rapid pulse rate change to detect an emotional event in the absence of physical effort such as patients trying to stand up or extend his/her arms. It has also been demonstrated that motion impacts the accuracy of wearable oximeter devices negatively [30].

This research goes beyond simply identifying and labeling emotions in real-time during ALS consultations. Its practical aim is to equip clinicians with immediate feedback during conversation. This feedback highlights specific moments that might raise concerns and warrant further exploration with the patient. Consequently, the provider can deepen their understanding of what might be sensitive for the patient and tailor their approach accordingly [2] [3].

The patients were selected to make sure that their speech was understandable [16] to use with NLP and represented a diverse population with no history of emotional sensitivity. An exclusion criterion was presence of severe depression or cognitive issues, underlying cardiac conditions as well as patients who already have feeding tubes or breathing assistance.

It is important to acknowledge that an early dementia diagnosis can be challenging, and some patients may fall on a spectrum of cognitive decline. Additionally, the term “dementia” often carries a negative connotation, which can discourage patients from seeking a specialist’s evaluation. Unexpectedly, patient 11 showed early signs of dementia during the study visit and we found that this patient was clearly an outlier for some of our metrics related to stress level.

This study aimed to provide clinicians with a systematic and unbiased dataset to support their daily work with ALS patients. We employed a set of validated digital tools previously tested in other contexts. Figure 8 presents a simplified conceptual diagram illustrating how these tools could theoretically be combined to advance the field of stress and depression assessment in patients experiencing various moods. This concept diagram might be used to interpret the psychiatry report in a simplified way. Upper triangle consists of observable outputs we can measure.

**Figure 8:**
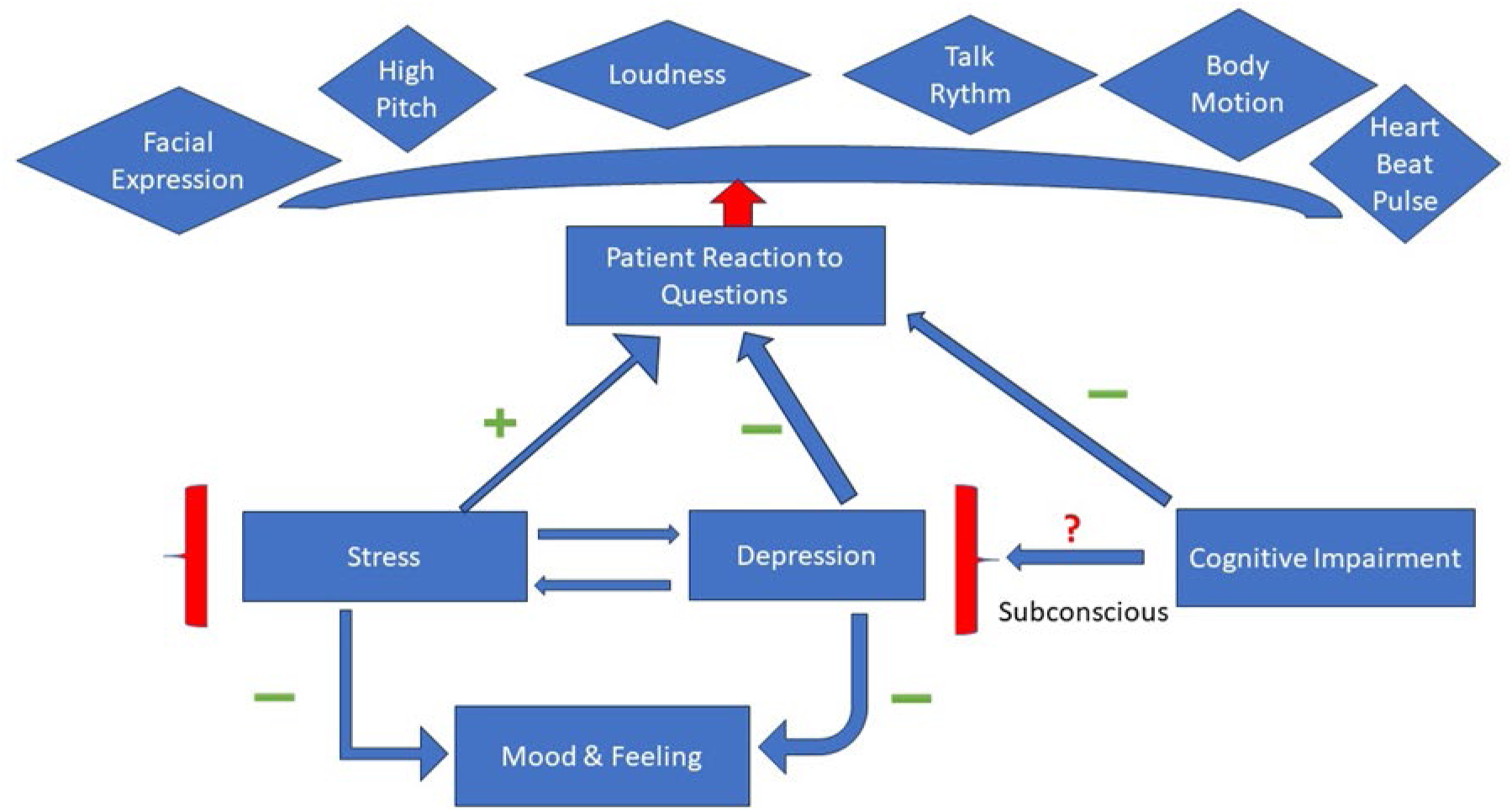
Conceptual diagram of the patient’s emotional state.

However, ALS patients often present atypical responses in these areas due to muscular weakness [31]. Therefore, this study focused on heart rate and natural language processing, assuming the patient’s speech remained intelligible.

Plus, and minus signs in Figure 8 companion the arrows to explicit the relationship. For example, increased stress promotes patient reaction intensity but depression does the opposite. Mood and feelings are reported by the patient and might be affected by both high stress and depression in a negative way. There is an interplay between stress and depression but not necessarily a bijective one. The subconscious impact of cognitive on stress and depression may persist with dementia.

Despite the inherent difficulty in measuring psychiatric characteristics with digital tools due to the lack of a universally agreed-upon definition of emotions (like the concept of “emotional arousal” used in this paper), a multi-modal approach combining voice analysis, body motion, facial expression, and physiological metrics offers the most comprehensive assessment.

The term “sentiment” in this context of NLP may be overly simplistic. This AI method relies on an annotated dictionary, which can miss the nuances of language and the semantic context that alters word meaning. Significant progress is needed to capture patient-specific feedback accurately. To address this, we implemented a hybrid approach combining emotional computing and NLP. This generates a concise report easily formatted by GLPs within minutes, assisting the multidisciplinary medical care team in clinical decision-making. The accuracy of the final report synthesis provided by a GLP assistant should be carefully reviewed by a clinician. Due to the rapid evolution of GLP technology, these systems cannot be entirely trusted, especially as model size increases, which can lead to deceptive outputs [32]. To mitigate these risks, we have simplified the information provided to the GLP software, focusing on patient concerns thanks to a combination of NLP and emotional computing techniques. However, as reported in our PCT [33], we plan to employ a variety of NLP methods to further enhance the quality and reliability of the generated reports with adequate post-processing. The ultimate goal is to provide real-time feedback to physicians, allowing them to tailor their approach and address underlying emotional concerns. Additionally, this can be particularly valuable in telehealth consultations where nonverbal cues are less readily available.

## 5. Conclusion

This study underscores the complexities of emotional assessment in ALS patients, highlighting the need for a multi-modal approach that accounts for the disease’s limitations. Future research should focus on longitudinal studies, delving deeper into the psychiatric aspects of ALS [34, 35, 36]. By integrating advanced natural language processing with physiological and behavioral data, we can potentially develop more comprehensive and accurate tools for emotional assessment, ultimately improving the quality of care for ALS patients and beyond.

## Author Contributions

Conceptualization, M.G, G. Ö., E.B.; methodology, M.G., Q.L.; software, M.G. Q.L.; validation: M.G., Q.L., G. Ö., V.G., H.J.K., E.B.; writing, review and editing: M.G., Q.L., G.Ö., V.G., H.J.K., E.B.

## Institutional Review Board Statement

The study was approved by the Institutional Review Board of the George Washington University (IRB# NCR235325).

## Informed Consent Statement

Informed consent was obtained from all subjects involved in the study.

## Data Availability Statement

The data presented in this study are available on request from the corresponding author due to patient privacy.

## Appendix A

Example below of a report generated automatically for Patient 1; notice that the length of the report is a parameter that can be set in our code.

**Report:** Assigning specific stress levels is subjective and can vary based on individual experiences and coping mechanisms. However, based on the information provided, we can make general assumptions about the potential stress levels associated with each concern:

***Swallowing and Choking Episodes:*** High stress. Difficulty swallowing and experiencing choking episodes can be emotionally distressing and may impact the patient’s daily life and sense of safety.

***Mobility and Walking Difficulty:*** High stress. The increasing stress related to mobility issues and bad balance can significantly impact the patient’s independence and quality of life, contributing to high stress levels.

***Breathing Perception:*** Moderate stress. While the patient expresses a concern about not feeling like they are getting enough air, the doctor’s assessment suggests good breathing results. The discrepancy between perception and medical evaluation may cause moderate stress.

***Medication Side Effects*** (Radicava): Moderate stress. Concerns about medication side effects, especially if they contribute to fatigue, can be moderately stressful. This stress level might increase if the fatigue affects daily functioning.

***Avoidance of Invasive Medical Interventions:*** Moderate stress. The patient expresses reluctance towards monitoring for choking episodes and a desire to avoid invasive interventions. The stress level is moderate, reflecting concerns about end-of-life decisions and potential medical procedures.

***Overall Health and Medical Review:*** Low to moderate stress. The general health review and medication discussion cover a broad range of topics but may not be an immediate source of stress for the patient. The stress level is lower compared to more specific health concerns.

It’s important to note that stress levels can vary based on individual perspectives, coping strategies, and the overall context of the patient’s health journey. Additionally, emotional responses may evolve over time as the patient receives more information and support from healthcare professionals and their support system.

## References

[1] Edwards WF, Malik S, Peters J, Chippendale I, Ravits J. Delivering Bad News in Amyotrophic Lateral Sclerosis: Proposal of Specific Technique ALS ALLOW. Neurol Clin Pract. 2021 Dec;11(6):521–526. doi: 10.1212/CPJ.0000000000000957. PMID: 34992959; PMCID: PMC8723938.

[2] Hogden A, Foley G, Henderson RD, James N, Aoun SM. Amyotrophic lateral sclerosis: improving care with a multidisciplinary approach. J Multidiscip Healthc. 2017 May 19;10:205–215. doi: 10.2147/JMDH.S134992. PMID: 28579792; PMCID: PMC5446964.

[3] Connolly S, Galvin M, Hardiman O. End-of-life management in patients with amyotrophic lateral sclerosis. Lancet Neurol. 2015 Apr;14(4):435–42. doi: 10.1016/S1474-4422(14)70221-2. Epub 2015 Feb 27. PMID: 25728958.

[4] Norris L, Que G, Bayat E. Psychiatric aspects of amyotrophic lateral sclerosis (ALS). Curr Psychiatry Rep. 2010 Jun;12(3):239–45. doi: 10.1007/s11920-010-0118-6. PMID: 20425287.

[5] Hadas Shahar, Hagit Hel-Or, Micro Expression Classification using Facial Color and Deep Learning Methods, The IEEE International Conference on Computer Vision (ICCV), 2019.

[6] Felipe Zago Canal, Tobias Rossi Müller, Jhennifer Cristine Matias, Gustavo Gino Scotton, Antonio Reis de Sa Junior, Eliane Pozzebon, Antonio Carlos Sobieranski,, A survey on facial emotion recognition techniques: A state-of-the-art literature review, Information Sciences, Volume 582, 2022, Pages 593–617, ISSN 0020-0255, 10.1016/j.ins.2021.10.005.

[7] Pavlovic, Vladimir I.; Sharma, Rajeev; Huang, Thomas S. (1997). “Visual Interpretation of Hand Gestures for Human–Computer Interaction: A Review” (PDF). IEEE Transactions on Pattern Analysis and Machine Intelligence. 19 (7): 677–695. doi:10.1109/34.598226. S2CID 7185733.

[8] Rosalind W Picard, Affective computing, MIT Press, 2000/7/24

[9] J. Yi, T. Nasukawa, R. Bunescu and W. Niblack, “Sentiment analyzer: extracting sentiments about a given topic using natural language processing techniques,” Third IEEE International Conference on Data Mining, Melbourne, FL, USA, 2003, pp. 427–434, doi: 10.1109/ICDM.2003.1250949.

[10] Julia Hirschberg and Christopher D. Manning, Advances in natural language processing, Science 349 (6245), pp261-266, 2015. DOI: 10.1126/science.aaa8685

[11] Guo, Jia. “Deep learning approach to text analysis for human emotion detection from big data” Journal of Intelligent Systems, vol. 31, no. 1, 2022, pp. 113–126. 10.1515/jisys-2022-0001

[12] Immordino-Yang MH, Yang XF, Damasio H. Cultural modes of expressing emotions influence how emotions are experienced. Emotion. 2016 Oct;16(7):1033–9. doi: 10.1037/emo0000201. Epub 2016 Jun 6. PMID: 27270077; PMCID: PMC5042821.

[13] Ford BQ, Mauss IB. Culture and emotion regulation. Curr Opin Psychol. 2015 Jun 1;3:1–5. doi: 10.1016/j.copsyc.2014.12.004. PMID: 25729757; PMCID: PMC4341898.

[14] Masrori P, Van Damme P. Amyotrophic lateral sclerosis: a clinical review. Eur J Neurol. 2020 Oct;27(10):1918–1929. doi: 10.1111/ene.14393. Epub 2020 Jul 7. PMID: 32526057; PMCID: PMC7540334.

[15] Zimmerman EK, Eslinger PJ, Simmons Z, Barrett AM. Emotional perception deficits in amyotrophic lateral sclerosis. Cogn Behav Neurol. 2007 Jun;20(2):79–82. doi: 10.1097/WNN.0b013e31804c700b. PMID: 17558250; PMCID: PMC1905862

[16] Eshghi, M., Yunusova, Y., Connaghan, K.P. et al. Rate of speech decline in individuals with amyotrophic lateral sclerosis. Sci Rep 12, 15713 (2022). 10.1038

[17] Helena E. A. Aho-Özhan, Jürgen Keller, Johanna Heimrath, Ingo Uttner, Jan Kassubek, Niels Birbaumer, Albert C. Ludolph, Dorothée Lulé, Perception of Emotional Facial Expressions in Amyotrophic Lateral Sclerosis (ALS) at Behavioural and Brain Metabolic Level, Published: October 14, 2016, 10.1371/journal.pone.0164655

[18] Jesse M. Cedarbaum, Nancy Stambler, Errol Malta, Cynthia Fuller, Dana Hilt, Barbara Thurmond, Arline Nakanishi, The ALSFRS-R: a revised ALS functional rating scale that incorporates assessments of respiratory function, Journal of the Neurological Sciences, Volume 169, Issues 1–2,1999, Pages 13–21, ISSN 0022-510X, 10.1016/S0022-510X(99)00210-5. Hutto, C.J. & Gilbert, E.E. (2014).

[19] AssemblyAI. Available online: https://www.assemblyai.com/

[20] Josiane Jauniaux, Marie-Hélène Tessier, Sophie Regueiro, Florian Chouchou, Alexis Fortin-Côté, Philip L. Jackson, Emotion regulation of others’ positive and negative emotions is related to distinct patterns of heart rate variability and situational empathy. PLoS ONE 15(12): e0244427. 2020 10.1371/journal.pone.0244427

[21] Wascher CAF. Heart rate as a measure of emotional arousal in evolutionary biology. Philos Trans R Soc Lond B Biol Sci. 2021 Aug 16;376(1831):20200479. doi: 10.1098/rstb.2020.0479. Epub 2021 Jun 28. PMID: 34176323; PMCID: PMC8237168.

[22] William H. Press, Saul A. Teukolsky, William T. Vetterling and Brian P. Flannery, Numerical Recipes in C, Cambridge University Press, 1992.

[23] Hutto, C.J. & Gilbert, E.E. (2014). VADER: A Parsimonious Rule-based Model for Sentiment Analysis of Social Media Text. Eighth International Conference on Weblogs and Social Media (ICWSM-14). Ann Arbor, MI, June 2014.

[24] Pols J, Limburg S. A Matter of Taste? Quality of Life in Day-to-Day Living with ALS and a Feeding Tube. Cult Med Psychiatry. 2016 Sep;40(3):361–82. doi: 10.1007/s11013-015-9479-y. PMID: 26547696; PMCID: PMC4945678.

[25] Nichols NL, Van Dyke J, Nashold L, Satriotomo I, Suzuki M, Mitchell GS. Ventilatory control in ALS. Respir Physiol Neurobiol. 2013 Nov 1;189(2):429–37. doi: 10.1016/j.resp.2013.05.016. Epub 2013 May 18. PMID: 23692930; PMCID: PMC4361018.

[26] Pagnini F, Rossi G, Lunetta C, Banfi P, Corbo M. Clinical psychology and amyotrophic lateral sclerosis. Front Psychol. 2010 Jul 21;1:33. doi: 10.3389/fpsyg.2010.00033. PMID: 21833203; PMCID: PMC3153752.

[27] Derek W. Johnston and Pavlos Anastasiades, The Relationship Between Heart Rate and Mood in Real Life, Journal of Psychosomatic Research, Vol. 34. No. I, pp. 21–27. 1990.

[28] Köbele R, Koschke M, Schulz S, Wagner G, Yeragani S, Ramachandraiah CT, Voss A, Yeragani VK, Bär KJ. The influence of negative mood on heart rate complexity measures and baroreflex sensitivity in healthy subjects. Indian J Psychiatry. 2010 Jan;52(1):42–7. doi: 10.4103/0019-5545.58894. PMID: 20174517; PMCID: PMC2824980.

[29] Yan Wu, Ruolei Gu, Qiwei Yang, Yue-jia Luo, How Do Amusement, Anger and Fear Influence Heart Rate and Heart Rate Variability? Front. Neurosci.,18 October 2019 Sec. Decision Neuroscience Volume 13 - 2019 | 10.3389/fnins.2019.01131

[30] Steven J. Barker, PhD, MD; Nitin K. Shah, MD, Effects of Motion on the Performance of Pulse Oximeters in Volunteers, Clinical Science, Anesthesiology October 1996, Vol. 85, 774– 781.

[31] Carelli L, Solca F, Tagini S, Torre S, Verde F, Ticozzi N, Consonni M, Ferrucci R, Pravettoni G, Poletti B, Silani V. Emotional Processing and Experience in Amyotrophic Lateral Sclerosis: A Systematic and Critical Review. Brain Sci. 2021 Oct 15;11(10):1356. doi: 10.3390/brainsci11101356. PMID: 34679420; PMCID: PMC8534224.

[32] Thilo Hagendorff, Deception abilities emerged in large language models, PNAS June 4, 2024, https://orcid.org/0000-0002-4633-2153

[33] Garbey, et al, Self-Assessment Neurological Health Care System, PCT/US2024/041422, August 8^th^2024.

[34] Zigmond AS, Snaith RP. The hospital anxiety and depression scale. Acta Psychiatr Scand. 1983 Jun;67(6):361–70. doi: 10.1111/j.1600-0447.1983.tb09716.x. PMID: 6880820.

[35] Birgitta Jakobsson Larsson, Karin Nordin, Ingela Nygren, Symptoms of anxiety and depression in patients with amyotrophic lateral sclerosis and their relatives during the disease trajectory, Journal of the Neurological Sciences, Volume 455, 2023, 10.1016/j.jns.2023.122780.

[36] Benbrika S, Desgranges B, Eustache F, Viader F. Cognitive, Emotional and Psychological Manifestations in Amyotrophic Lateral Sclerosis at Baseline and Overtime: A Review. Front Neurosci. 2019 Sep 10;13:951. doi: 10.3389/fnins.2019.00951. PMID: 31551700; PMCID: PMC6746914.

